# Menopause timing, epigenetic aging and blood-cell traits

**DOI:** 10.64898/2026.10.05.26364790

**Authors:** Xin Fang, Joel Schwartz

## Abstract

Menopause timing links reproductive lifespan with later-life health, motivating investigation of its molecular and hematological correlates. We combined Generation Scotland analyses of women sampled at or after reported menopause (n = 1,444) and initially menstruating women followed for GP-ascertained menopause onset (n = 1,725; 251 events) with bidirectional and multivariable Mendelian randomization. Higher age-adjusted GrimAge V1, GrimAge V2 and DunedinPACE corresponded to 0.34–0.40 years earlier menopause per SD in the retrospective cohort at nominal P < 0.05. Prospective associations were nonsignificant, and cohort and primary bidirectional aging-related genetic associations did not survive multiple-testing correction. Genetically predicted later menopause was associated with higher neutrophil percentage and count and lower lymphocyte percentage in women; these associations and female–male contrasts survived correction. CRP conditional on menopause timing was associated with higher GrimAge and PhenoAge acceleration and shorter leukocyte telomere length after correction. None of 36 estimated inflammatory indirect effects reached P < 0.05. These findings highlight blood-cell composition as a prominent correlate of reproductive timing and identify selected methylation-aging signatures for further study. Together, they position reproductive–hematological relationships as a specific avenue for understanding the connection between menopause and systemic aging.

## Introduction

Reproductive aging offers a clinically observable window into processes relevant to health across the life course. Earlier natural menopause is associated with higher subsequent cardiovascular disease risk [1], while genetic studies implicate genome-maintenance pathways in the timing of ovarian aging [2] [3]. Characterizing the blood-based phenotypes associated with menopause timing can help connect this reproductive milestone with the wider biology of aging.

DNA methylation provides measures of chronological age, mortality-related risk and the pace of physiological aging [4] [5] [6] [7]. Earlier menopause has been associated with greater blood epigenetic age acceleration and higher GrimAge [8] [6]. Recent NHANES analyses reported associations involving GrimAge, reproductive profiles and DunedinPoAm, or last-period age and VidalBralo age acceleration [9] [10] [11]. The Health and Retirement Study further linked early menopause without hysterectomy to a composite epigenetic-aging measure [12]. These findings establish menopause timing as a relevant reproductive phenotype for molecular-aging research and motivate comparison across measures with different biological targets.

Sampling relative to menopause determines the question that these measures address. Methylation measured after menopause characterizes differences between women with different reproductive histories; measurement before menopause permits assessment of subsequent onset. In CARDIA, a prospective GrimAge association attenuated after broader covariate adjustment [13]. Bidirectional genetic analyses offer a complementary approach to directionality [14], and broader reproductive-trait studies have combined observational and genetic evidence [15]. Bringing these designs together can clarify where menopause timing is reflected in aging-related phenotypes.

Blood-cell composition and inflammation provide a biological context for this comparison. A menopause phenome-wide genetic study identified blood-cell traits as an important area for follow-up [16]. Longitudinal human studies have documented leukocyte trajectories around the final menstrual period and changes in gut-barrier and microbial-translocation-related immune markers across the transition [17] [18]. In parallel, health-related methylation measures capture inflammatory information: DNAm PhenoAge is associated with inflammatory transcriptional signatures, and GrimAge V2 includes a methylation surrogate of C-reactive protein (CRP) [5] [19]. These observations motivate examination of both measured blood-cell traits and specific inflammatory pathways.

Here, we examined menopause timing in relation to epigenetic aging using complementary Generation Scotland cohorts, assessed genetic directions of association, and investigated blood-cell and inflammatory phenotypes. The study addresses three questions: how methylation measures relate to menopause before and after its occurrence; how genetic predictors of menopause timing relate to circulating blood-cell traits in women and men; and whether inflammatory biomarkers account for an indirect association with aging-related outcomes.

## Results

### Cohorts and analytical scope

The retrospective cohort included 1,444 women from 1,118 family clusters. Mean baseline age was 58.9 years (SD 8.6), and mean reported age at natural menopause (ANM) was 49.0 years (SD 5.1). The prospective cohort included 1,725 initially menstruating women from 1,374 families, with a mean baseline age of 36.7 years (SD 9.9). During 18,880 person-years of stored follow-up, 251 women had a GP-ascertained menopause onset and 1,474 were censored; median follow-up was 11 years (interquartile range 10–13) (Table 1; Supplementary Fig. S1).

**Table 1.**
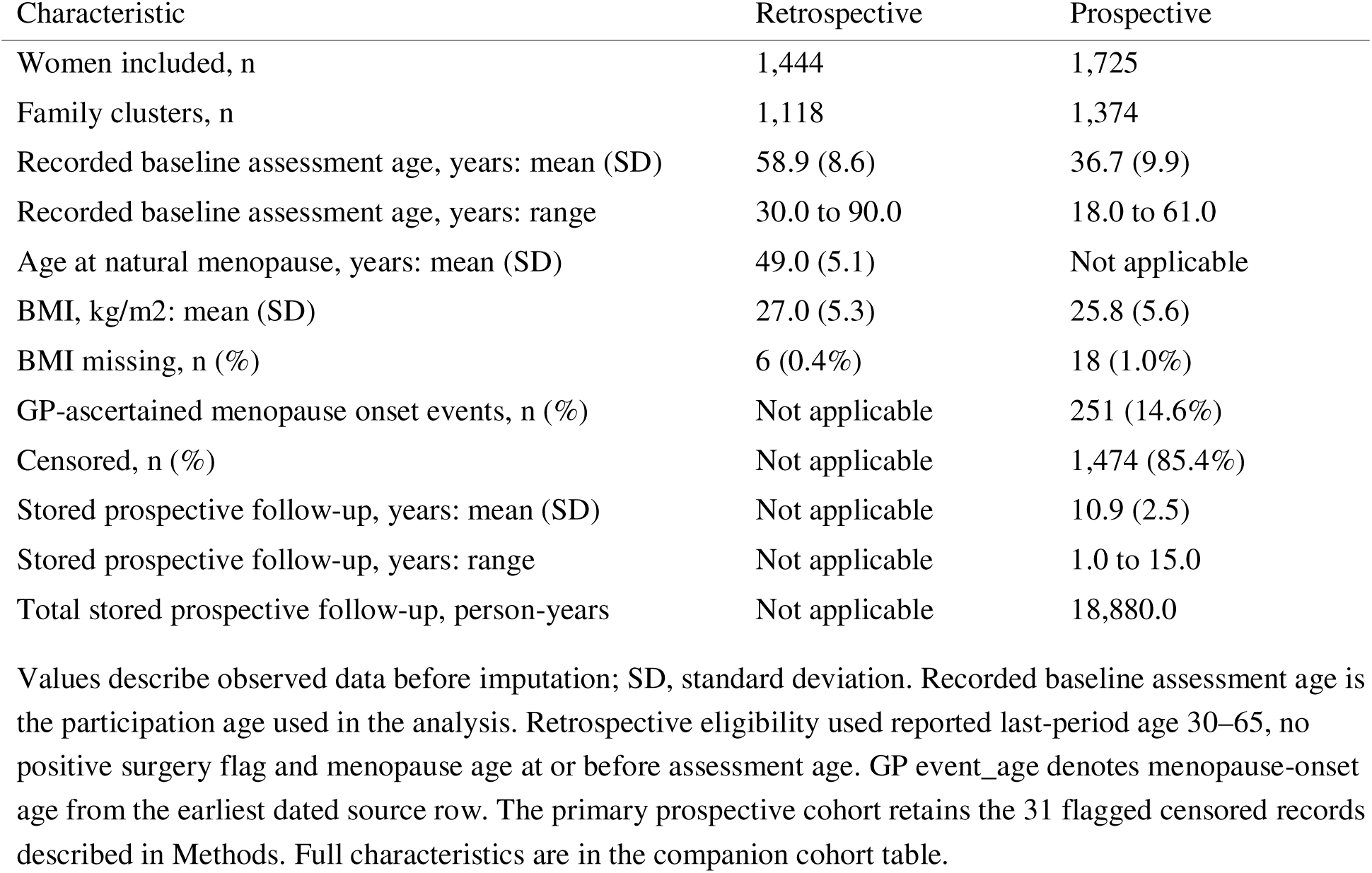
Primary Generation Scotland cohort characteristics.

Cohort analyses covered thirteen methylation markers. Ten conventional measures are displayed in the main results; three principal-component (PC) implementations are reported as supporting analyses. All shared the same imputation and adjustment procedures, and the complete 26-test marker-by-cohort correction family was retained. Genetic analyses included four epigenetic-clock phenotypes and five further aging-related datasets. Of 95 planned univariable pairs, 73 were estimable; 36 of 50 planned multivariable models and corresponding indirect effects were available (Supplementary Table S1; Supplementary Data 2). Throughout, P values are two-sided; nominal associations have P < 0.05, and q < 0.05 denotes support after Benjamini–Hochberg correction within the stated family.

### Methylation-aging associations and genetic direction

In the retrospective cohort, higher age-adjusted GrimAge V1, GrimAge V2 and DunedinPACE were associated with earlier ANM at nominal P < 0.05. Per SD higher score, ANM was 0.395 years earlier for GrimAge V1 (95% CI −0.691 to −0.099; P = 0.00902), 0.366 years earlier for GrimAge V2 (−0.666 to −0.065; P = 0.0171), and 0.343 years earlier for DunedinPACE (−0.629 to −0.057; P = 0.019). Reversed DNAm telomere length (β = −0.349; −0.609 to −0.090; P = 0.0084) and Zhang mortality (β = −0.444; −0.744 to −0.143; P = 0.00386) showed the same direction. None of the 26 cohort associations met q < 0.05; q values for these five retrospective associations ranged from 0.0782 to 0.0986 (Fig. 1; Table 2; Supplementary Table S4).

**Figure 1.**
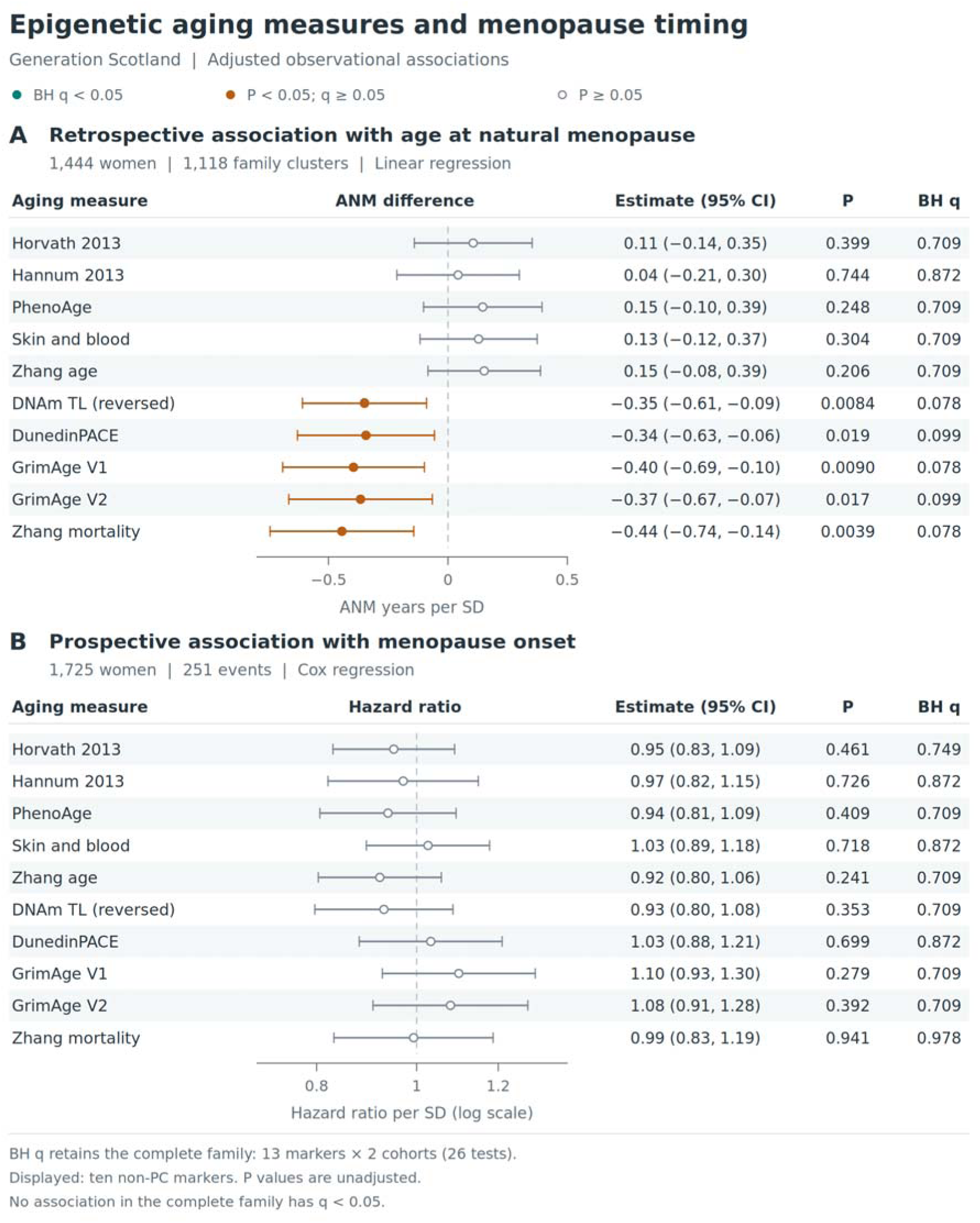
Associations of epigenetic aging measures with menopause timing in Generation Scotland. Figures report nominal two-sided P values and Benjamini–Hochberg (BH) q values. Teal denotes q < 0.05, amber denotes P < 0.05 with q ≥ 0.05, and open slate points denote P ≥ 0.05. Correction families are specified below. ANM, age at natural menopause; CI, confidence interval; MR, Mendelian randomization. **A**, Retrospective associations in 1,444 women from 1,118 family clusters, expressed as differences in ANM years per SD higher age-adjusted methylation marker. **B**, Prospective associations in 1,725 women from 1,374 family clusters, including 251 subsequent GP-ascertained menopause onsets, expressed as hazard ratios per SD on a logarithmic axis. Points and horizontal lines represent pooled estimates and 95% CIs. The ten conventional markers are shown in the same fixed order in both panels. The three supporting PC clocks are displayed in Supplementary Fig. S3. Scores were residualized on a three-degree-of-freedom baseline-age spline and standardized within cohort; DNAm telomere length was reversed so higher values denote shorter predicted telomeres. Separate linear or attained-age Cox models adjusted for age, BMI, smoking proxy, education, six estimated cell fractions and methylation batch, using the same twenty-imputation run and family-robust uncertainty. The prospective analysis used delayed entry and batch-stratified baseline hazards. BH q values retain all thirteen markers across both cohorts (26 tests), including the supporting PC clocks; none has q < 0.05. Proportional-hazards diagnostics and sensitivity analyses are reported in Supplementary Table S5 and Methods S8.

**Table 2.** Associations of conventional methylation markers with menopause timing.

| Marker | Retrospective $\beta$ (95% CI) | P | Prospective HR (95% CI) | P |
| --- | --- | --- | --- | --- |
| Horvath 2013 | 0.106 (−0.141, 0.353) | 0.399 | 0.950 (0.830, 1.088) | 0.461 |
| Hannum 2013 | 0.043 (−0.214, 0.299) | 0.744 | 0.971 (0.821, 1.148) | 0.726 |
| PhenoAge | 0.146 (−0.102, 0.394) | 0.248 | 0.938 (0.806, 1.092) | 0.409 |
| Skin and Blood | 0.129 (−0.117, 0.374) | 0.304 | 1.026 (0.894, 1.177) | 0.718 |
| Zhang age | 0.152 (−0.084, 0.388) | 0.206 | 0.921 (0.803, 1.057) | 0.241 |
| DNAm telomere length (reversed) | −0.349 (−0.609, −0.090) | 0.0084 | 0.930 (0.797, 1.084) | 0.353 |
| DunedinPACE | −0.343 (−0.629, −0.057) | 0.019 | 1.032 (0.880, 1.211) | 0.699 |
| GrimAge V1 | −0.395 (−0.691, −0.099) | 0.00902 | 1.099 (0.926, 1.303) | 0.279 |
| GrimAge V2 | −0.366 (−0.666, −0.065) | 0.0171 | 1.078 (0.907, 1.282) | 0.392 |
| Zhang mortality | −0.444 (−0.744, −0.143) | 0.00386 | 0.993 (0.832, 1.186) | 0.941 |

Prospective hazard ratios per SD were 1.099 for GrimAge V1 (95% CI 0.926–1.303; P = 0.279), 1.078 for GrimAge V2 (0.907–1.282; P = 0.392), and 1.032 for DunedinPACE (0.880–1.211; P = 0.699). All prospective associations had P ≥ 0.05. GrimAge V2 retrospective sensitivity coefficients remained negative across the examined adjustments and eligibility restrictions (−0.461 to −0.278 years per SD). Excluding the 31 flagged prospective censored records retained all 251 events and yielded a V2 hazard ratio of 1.068 (0.899–1.268). Proportional-hazards departures prompted age-varying sensitivity analyses, reported with the complete diagnostics in Supplementary Table S5 and Methods S8. Supporting PC-clock associations also had P ≥ 0.05 in both cohorts (Supplementary Table S5; Supplementary Fig. S3).

Forward MR used 153–155 ANM instruments for the four clock outcomes. None of the forward clock estimates reached P < 0.05 (Table 3). In reverse MR, higher genetically predicted original GrimAge acceleration was nominally associated with earlier ANM (β = −0.1287 years per source GWAS unit; 95% CI −0.2565 to −0.00095; P = 0.0483; q = 0.483). This estimate used four instruments, showed heterogeneity (I² = 71.5%), and had weighted median, weighted mode and MR-Egger P values ≥ 0.05. No primary forward or reverse aging-related MR association met its correction threshold (Fig. 2; Supplementary Table S2).

**Figure 2.**
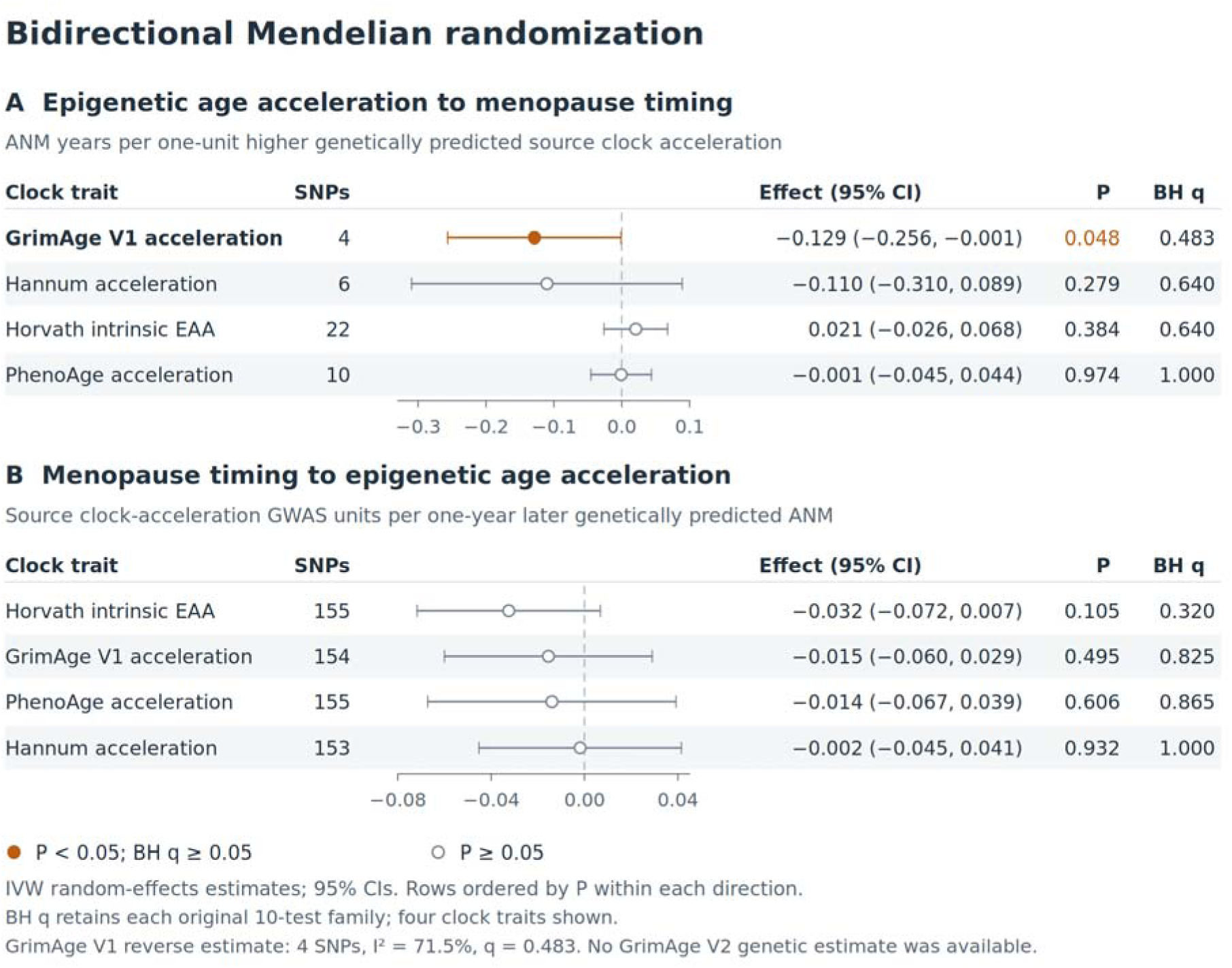
Bidirectional genetic associations of epigenetic aging and menopause timing. Figures report nominal two-sided P values and Benjamini–Hochberg (BH) q values. Teal denotes q < 0.05, amber denotes P < 0.05 with q ≥ 0.05, and open slate points denote P ≥ 0.05. Correction families are specified below. ANM, age at natural menopause; CI, confidence interval; MR, Mendelian randomization. **A**, Reverse MR from the four epigenetic aging traits to ANM. **B**, Forward MR from ANM to the four epigenetic aging traits. Within each direction, rows are ordered by increasing unadjusted P. Points and horizontal lines indicate primary random-effects IVW estimates and 95% CIs; the number of retained variants, unadjusted P and original-family BH q accompany each estimate. Reverse coefficients are ANM years per source epigenetic-age-acceleration GWAS unit; forward coefficients are source epigenetic-age-acceleration GWAS units per ANM year. The two directions have separate axes and source-specific units. BH correction uses the original ten-test family for each direction. The nominal reverse GrimAge association (P = 0.0483; q = 0.483) uses four variants and has I² = 71.5%; weighted-median, weighted-mode and MR-Egger sensitivity estimates had P ≥ 0.05. All forward clock estimates are nonsignificant. GrimAge refers to the original V1 phenotype. Generation Scotland contributed to the clock GWAS, creating participant overlap between the cohort and genetic evidence.

**Table 3.** Bidirectional MR of ANM and four epigenetic age-acceleration traits.

| Direction | Trait | SNPs | $\beta$ (95% CI) | P |
| --- | --- | --- | --- | --- |
| ANM → clock | GrimAge V1 acceleration | 154 | -0.01550 (-0.05999, 0.02899) | 0.4948 |
| ANM → clock | PhenoAge acceleration | 155 | -0.01401 (-0.06716, 0.03915) | 0.6055 |
| ANM → clock | Hannum acceleration | 153 | -0.00188 (-0.04524, 0.04148) | 0.9322 |
| ANM → clock | Horvath intrinsic EAA | 155 | -0.03246 (-0.07169, 0.00677) | 0.1048 |
| Clock → ANM | GrimAge V1 acceleration | 4 | -0.12872 (-0.25649, -0.00095) | 0.0483 |
| Clock → ANM | PhenoAge acceleration | 10 | -0.00074 (-0.04528, 0.04380) | 0.9742 |
| Clock → ANM | Hannum acceleration | 6 | -0.11025 (-0.30966, 0.08916) | 0.2785 |
| Clock → ANM | Horvath intrinsic EAA | 22 | 0.02081 (-0.02602, 0.06764) | 0.3837 |

### Sex-specific associations with measured blood-cell traits

Genetically predicted later ANM was associated with higher neutrophil percentage (β = 0.0204; 95% CI 0.0139–0.0268; P = 5.41 × 10⁻¹⁰), higher neutrophil count (β = 0.0205; 0.0112–0.0298; P = 1.65 × 10⁻⁵), and lower lymphocyte percentage (β = −0.0218; −0.0282 to −0.0153; P = 3.22 × 10⁻¹¹) in women. Effects are in source rank-normalized units per genetically predicted ANM year. All three IVW associations retained q < 0.05, with support across weighted median, weighted mode and MR-Egger estimates (Fig. 3; Supplementary Table S3).

**Figure 3.**
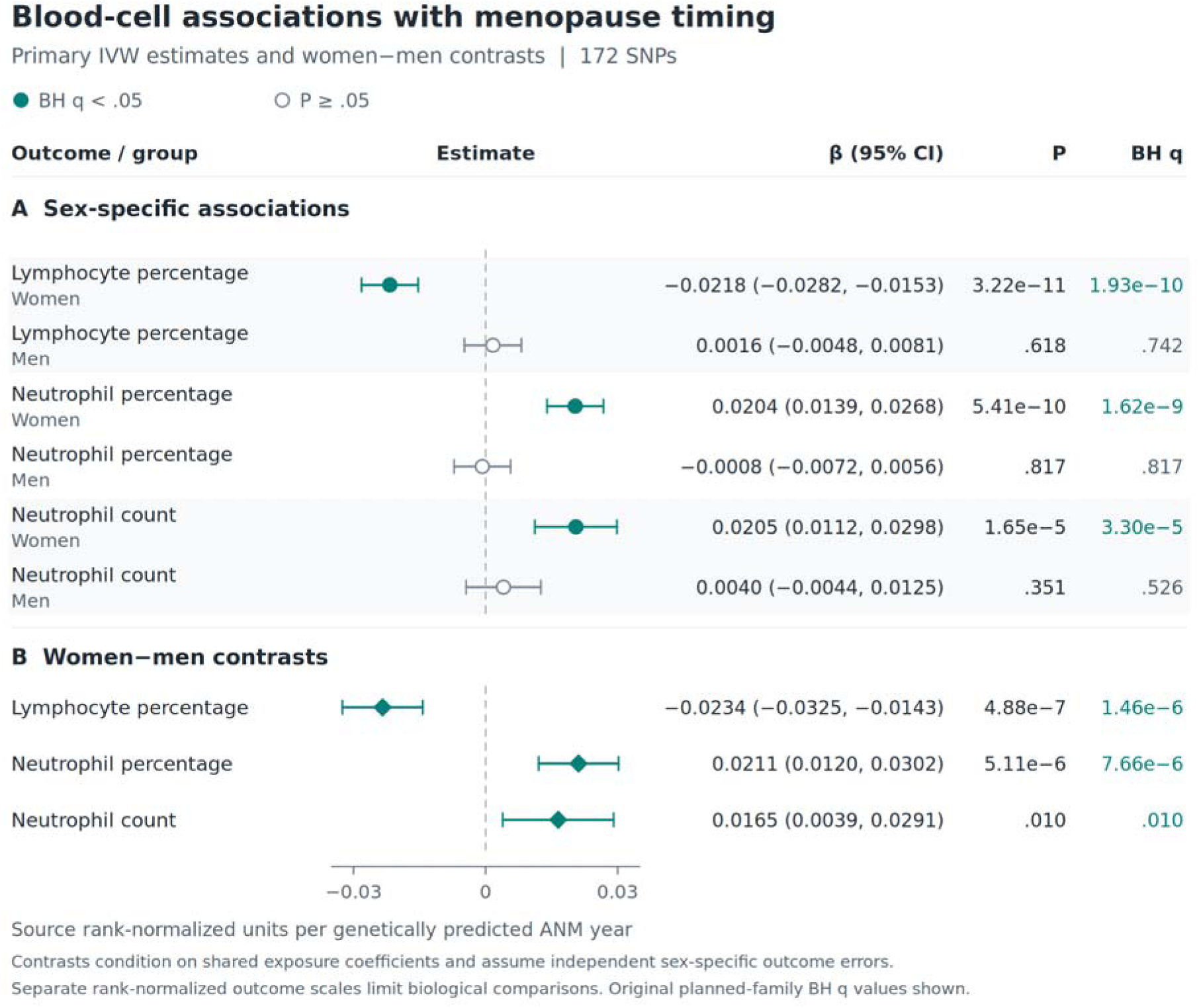
Sex-specific blood-cell associations with genetically predicted menopause timing. Figures report nominal two-sided P values and Benjamini–Hochberg (BH) q values. Teal denotes q < 0.05, amber denotes P < 0.05 with q ≥ 0.05, and open slate points denote P ≥ 0.05. Correction families are specified below. ANM, age at natural menopause; CI, confidence interval; MR, Mendelian randomization. **A**, Primary inverse-variance-weighted (IVW) estimates for genetically predicted later ANM and measured lymphocyte percentage, neutrophil percentage and neutrophil count in women and men. **B**, Corresponding women-minus-men coefficient contrasts. All estimates use 172 retained ANM instruments. Traits are ordered by increasing female IVW P; women precede the corresponding male comparison. Circles denote sex-specific estimates and diamonds denote contrasts; horizontal lines are 95% CIs. Units are source rank-normalized blood-cell trait units per genetically predicted ANM year. Original planned-family BH q values are shown: six sex-specific IVW tests and three sex contrasts are corrected in their respective families. Contrasts condition on the shared exposure coefficients and assume independent sex-specific outcome errors, without overlap-covariance correction. They retain the source-specific rank-normalized scales. Male IVW estimates are nonsignificant, although the male neutrophil-count weighted-median sensitivity estimate had P = 0.0320 and q = 0.0480. These analyses concern measured blood-cell abundance and composition; the mediation analyses use separate outcomes.

Female–male contrasts supported sex differences for neutrophil percentage (P = 5.11 × 10⁻⁶), neutrophil count (P = 0.0104) and lymphocyte percentage (P = 4.88 × 10⁻⁷), each with q < 0.05 under the stated covariance assumptions. Male IVW estimates included zero; the male neutrophil-count weighted median estimate had P = 0.0320 and q = 0.0480. These comparisons characterize sex-dependent genetic associations with measured cell abundance and composition (Supplementary Fig. S2).

### Inflammatory associations and indirect pathways

CRP conditional on ANM was associated with higher original GrimAge acceleration (β = 0.290; 95% CI 0.123–0.457; P = 0.000648; q = 0.0162), higher PhenoAge acceleration (β = 0.327; 0.116–0.538; P = 0.00236; q = 0.0393), and shorter Codd leukocyte telomere length (β = −0.0363; −0.0558 to −0.0168; P = 0.000268; q = 0.0134). Coefficients are in the corresponding outcome GWAS units per unit higher natural-log CRP concentration. These were the three conditional biomarker associations that met q < 0.05; the complete set of 36 coefficients is shown in Fig. 4. No conditional ANM coefficient met its separate correction threshold.

**Figure 4.**
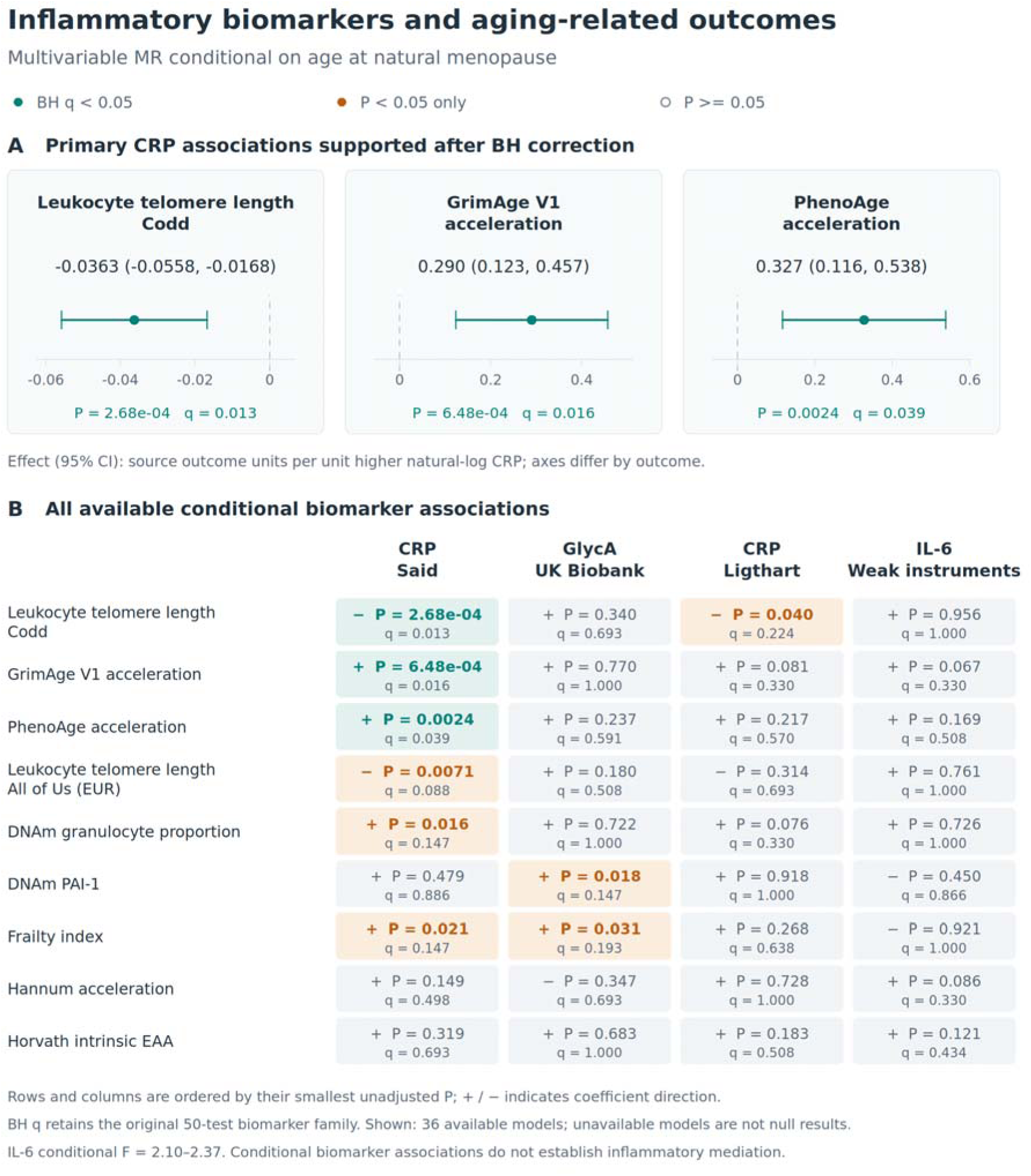
Conditional associations of inflammatory biomarkers with aging-related outcomes. Figures report nominal two-sided P values and Benjamini–Hochberg (BH) q values. Teal denotes q < 0.05, amber denotes P < 0.05 with q ≥ 0.05, and open slate points denote P ≥ 0.05. Correction families are specified below. ANM, age at natural menopause; CI, confidence interval; MR, Mendelian randomization. Multivariable MR jointly models ANM and each inflammatory biomarker. **A**, The three primary-source CRP coefficients supported at BH q < 0.05, presented by increasing unadjusted P. Points and horizontal lines indicate conditional coefficients and 95% CIs. Each panel has its own axis and outcome GWAS units per unit higher natural-log CRP concentration. **B**, All 36 available conditional biomarker coefficients across nine aging-related datasets and four biomarker sources. Each cell displays the coefficient sign, nominal P and original BH q; cell colors encode statistical support. Rows and columns are ordered by their minimum unadjusted P. BH correction retains the original 50-test biomarker-coefficient family, including unavailable planned tests. CRP, C-reactive protein; GlycA, glycoprotein acetyls; IL-6, interleukin-6; EAA, epigenetic age acceleration; PAI-1, plasminogen activator inhibitor-1; EUR, European ancestry. Conditional IL-6 instruments are weak (F = 2.10–2.37). The corresponding indirect estimates are shown in Figure 5. EstBB GlycA and FinnGen frailty models were unavailable.

The primary ANM-to-CRP IVW estimate was −0.00191 (95% CI −0.00916 to 0.00534; P = 0.606). Weighted median and mode estimates were negative and retained q < 0.05, with IVW heterogeneity of I² = 88.3%. The primary ANM-to-IL-6, GlycA and alternative-source CRP estimates had P ≥ 0.05. None of the 36 indirect products reached P < 0.05 (all P ≥ 0.285; all q = 1). For ANM–CRP–GrimAge, the indirect estimate was −0.00055 source GrimAge units per ANM year (95% CI −0.00268 to 0.00157; P = 0.610) (Fig. 5). Conditional F statistics were 112–145 for primary CRP, 95–98 for alternative CRP and 23–25 for GlycA; IL-6 instruments were weak conditionally (F = 2.10–2.37). All component estimates and diagnostics are provided in Supplementary Data 2 and Methods S10.

**Figure 5.**
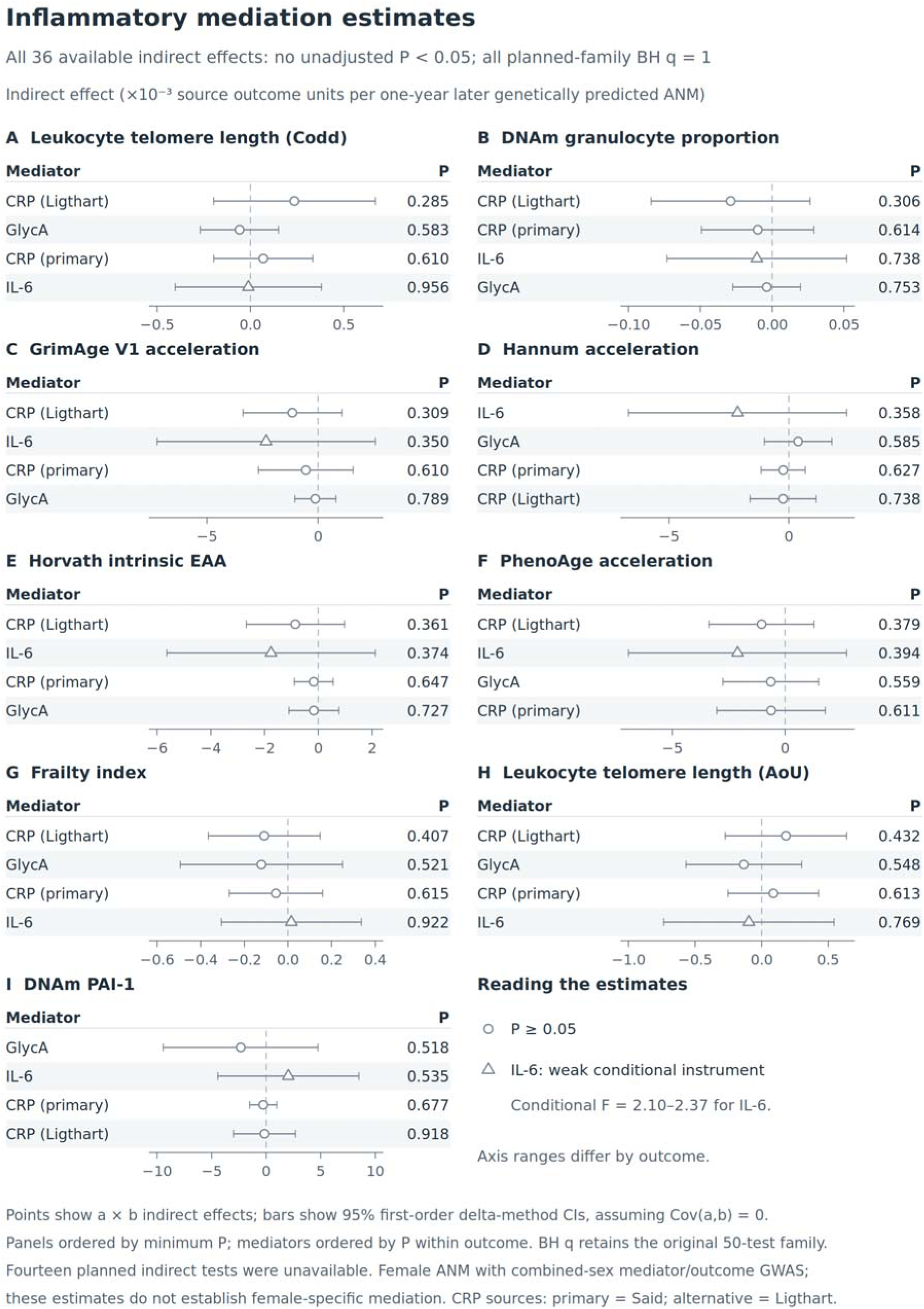
Indirect associations of menopause timing with aging-related outcomes through inflammatory biomarkers. Figures report nominal two-sided P values and Benjamini–Hochberg (BH) q values. Teal denotes q < 0.05, amber denotes P < 0.05 with q ≥ 0.05, and open slate points denote P ≥ 0.05. Correction families are specified below. ANM, age at natural menopause; CI, confidence interval; MR, Mendelian randomization. All 36 available ANM–biomarker–outcome indirect estimates are shown. Each outcome has a separate horizontal scale; displayed axis values are in units of 10⁻³ source outcome GWAS units per ANM year (multiply axis values by 0.001 to recover source units). Points and horizontal lines indicate the product a × b and 95% delta-method CIs, where a is the primary IVW ANM-to-biomarker coefficient and b is the biomarker coefficient conditional on ANM. Both paths use the same biomarker source. Outcomes are ordered by the smallest unadjusted P among their four mediator sources, and mediator rows are ordered by increasing unadjusted P within outcome, as documented in the plotted source table. Triangles mark IL-6 estimates with weak conditional instruments (F < 10); circles mark the other estimates. All P values are at least 0.285 and all original-family BH q values equal 1. The 50-test indirect-effect correction family retains the 14 unavailable planned models. Delta-method uncertainty assumes zero covariance between a and b, and conditional strength calculations assume zero cross-exposure sampling covariance. These covariance assumptions are subject to participant overlap across GWAS. The outcomes differ from the measured sex-specific blood-cell traits in Figure 3.

## Discussion

Our analyses identify blood-cell composition as a prominent feature of the relationship between reproductive timing and systemic aging. Later genetically predicted menopause was associated with higher neutrophil percentage and count and lower lymphocyte percentage in women, with support across estimators and in female–male contrasts after multiple-testing correction. The cohort analyses additionally identified nominal retrospective associations involving GrimAge and DunedinPACE, while CRP was associated with selected aging outcomes conditional on menopause timing. The study brings these findings together across sampling periods and biological measures, giving reproductive–hematological relationships a specific place in the study of women’s aging.

The retrospective methylation findings extend the evidence connecting reproductive history with health-related aging signatures. GrimAge was developed from mortality-related methylation surrogates, whereas DunedinPACE was trained against longitudinal physiological change [6] [7]. Their nominal associations with earlier menopause are consistent with a relationship involving these dimensions of aging. Earlier studies reported menopause associations with blood epigenetic age and GrimAge [8] [6], and recent NHANES and Health and Retirement Study analyses add context across individual and composite measures [9] [12]. Related NHANES analyses examined different reproductive contrasts and reported DunedinPoAm or VidalBralo associations [10] [11]. Several of these reports use overlapping participants; collectively, they motivate attention to the biological target of each measure and the reproductive phenotype being studied. In our data, the retrospective associations remain exploratory after correction.

The prospective and genetic analyses add temporal and directional information. Baseline methylation scores were not associated with subsequent menopause onset at P < 0.05, and the prospective GrimAge estimates were modest and imprecise. CARDIA similarly reported attenuation of its baseline GrimAge association after broader adjustment [13]. Forward clock MR estimates were compatible with earlier bidirectional analyses [14]; the nominal reverse GrimAge result provides a direction for follow-up within broader reproductive-aging research [15]. The contribution of these designs is to locate the observed methylation pattern relative to menopause: the present evidence concerns differences among women sampled at or after menopause more than prediction of subsequent onset. Repeated samples across the transition would connect these between-person associations to within-person trajectories.

The blood-cell analyses provide the clearest genetic connection with menopause timing. They extend a menopause phenome scan that identified cell composition as an important follow-up area [16]. Support across estimators, together with female–male contrasts, strengthens the case for studying the relationship in a sex-specific framework. Longitudinal data from Kailuan showed distinct leukocyte trajectories around the final menstrual period, including declining neutrophil counts over part of the transition [17]. SWAN recently documented increases in gut-barrier and microbial-translocation-related immune markers around menopause [18]. These studies address changes during the transition, while our genetic analyses address variation in its timing. Together, they establish a concrete rationale for integrating reproductive histories with circulating cell measurements. Cell composition is the phenotype supported here; activation states and tissue-specific immune processes are questions for subsequent mechanistic work.

The inflammatory analyses identify a complementary association between CRP-related variation and aging phenotypes. CRP conditional on ANM was associated with GrimAge, PhenoAge and leukocyte telomere length after correction, extending previous genetic work on these outcomes [20] [21]. The full indirect products did not establish mediation through the tested biomarkers. This separates the observed CRP–aging relationship from evidence for a menopause–inflammation–aging pathway and gives future studies a more specific target: measuring reproductive timing, cellular composition and inflammatory profiles jointly. The value of examining several biological systems is also supported by metabolomic work linking menopause duration with clinical aging measures [22].

The study’s strengths are its complementary cohort designs, consistent comparison of methylation measures, adjustment for family relatedness and integration of directional, sex-specific and pathway analyses. Its scope is defined by a single methylation assessment, 251 prospective onsets and operational menopause definitions with incomplete surgical histories and individual linkage coverage. Calendar-year censoring and proportional-hazards departures qualify the prospective estimates. Residual confounding, including hormone-therapy and parity information, and adjustment for covariates measured after menopause remain relevant. Missing CpGs were reference-imputed, and delivery-specific normalization and cell-estimation records are incomplete. Shared SGS and UK Biobank contributions, sparse reverse-clock instruments, weak conditional IL-6 instruments and unestimated sampling covariance constrain causal interpretation [23] [24]. Predominantly European genetic sources limit generalizability. The full estimates, source definitions and sensitivities are retained in the supplementary material.

Menopause timing is a useful phenotype for connecting reproductive lifespan with systemic biology. The clearest findings in this study concern sex-dependent blood-cell associations, accompanied by exploratory retrospective methylation-aging signals and CRP associations with selected aging outcomes. These results make reproductive–hematological relationships a focused priority for studies of the biological processes underlying women’s aging.

## Methods

### Design and participants

We combined Generation Scotland cohort analyses with bidirectional and multivariable MR. Analyses were exploratory and not preregistered. Reporting followed STROBE-MR principles [25]. Supplementary Methods S6–S11 provide the detailed analytical and provenance records.

Generation Scotland recruited participants during 2006–2011, using a pre-clinic questionnaire followed by clinic blood collection [26] [27]. The analysis used recorded baseline participation age; participant-specific questionnaire and blood dates were unavailable. Retrospective eligibility required reported last-period age 30–65 years, no ongoing menstruation and no positive hysterectomy or ovary-removal response. Missing status was retained when a qualifying age was available, and missing surgery answers denoted no known surgery. ANM was operationalized from cessation age without independent confirmation of twelve months of amenorrhea. The primary cohort required ANM at or before assessment age, retaining 1,444 of 1,447 eligible women.

Prospective eligibility required ongoing menstruation, GP-linkage consent, no positive surgery flag and positive stored follow-up. The endpoint was supplied GP-ascertained menopause-onset age from the earliest linked row ordered by Date. Events required onset after entry; otherwise, exit age was 2020 minus year of birth. Individual death, deregistration and practice-coverage dates were not incorporated. Exact project coverage and the approved GP code list were unavailable in the supplied records. The cohort included 1,725 women and 251 events. A sensitivity excluded 31 flagged censored records. Source-specific status coding, missing-response counts and endpoint checks are detailed in Supplementary Methods S6.

### Methylation measures

Whole-blood methylation was measured with the Illumina MethylationEPIC array. The delivered matrix contained 831,733 CpGs and 18,858 samples, including 11,092 female samples. Published processing describes dasen normalization and separately or jointly normalized releases [27]; the exact delivered release and the estimator for seven supplied cell fractions remain unidentified. Supplied M values were converted to beta values where required. Models used six supplied cell fractions, omitting eosinophils.

Conventional measures were Horvath 2013, Hannum, DNAm PhenoAge, Skin and Blood, Zhang chronological age, DunedinPACE, GrimAge V1 and V2, DNAm telomere length and Zhang mortality [4] [28] [5] [29] [30] [7] [6] [19] [31] [32]. PC-Horvath, PC-Hannum and PC-PhenoAge were evaluated as supporting measures using published fixed projections and coefficients [33]. No clock was fitted to menopause outcomes. Missing required CpGs were reference-imputed; implementation, coverage and scoring details are in Supplementary Methods S6–S7. Within each cohort, scores were residualized on a three-degree-of-freedom natural cubic spline of baseline age and standardized to one SD. DNAm telomere length was reversed so higher scores indicated shorter predicted telomeres. Score construction and reference filling were treated as fixed for inference.

### Cohort models and supporting analyses

Separate retrospective linear regressions used ANM years as outcome with family-clustered HC1 standard errors. Prospective Cox models used attained age, delayed entry, Efron handling of ties, batch-stratified baseline hazards and family-clustered robust variance. Models adjusted for the baseline-age spline, BMI, log(1 + smoking proxy), education, and B-cell, CD4-T, CD8-T, monocyte, neutrophil and natural-killer-cell fractions; retrospective models included batch fixed effects. Smoking used pack-years, assigning zero to reported never-smokers. Hormone therapy and parity were not included in the adjustment set.

Missing BMI, smoking and education were handled with 20 chained-equation imputations, ten iterations and seed 20261003. Predictive mean matching was used for continuous covariates and multinomial regression for education. Imputation included all thirteen markers, covariates and outcome information, including event status and the delayed-entry cumulative hazard prospectively. Coefficients and family-robust variances were pooled using Rubin’s rules with t-based inference. Imputation was single-level. Complete-case, eligibility and adjustment sensitivities retained primary score standardization. Proportional-hazards diagnostics were assessed in each completed dataset; exploratory age-varying models split follow-up at every event age. Full specifications, pooling details and diagnostics are in Supplementary Methods S8.

### Genetic data and instruments

The primary ANM GWAS included up to 201,323 European women with natural menopause at ages 40–60 [2]. European-stratum, combined-sex GWAS supplied original GrimAge, Hannum, Horvath intrinsic and PhenoAge acceleration [34]. Additional outcomes were DNAm granulocyte proportion, DNAm PAI-1, frailty and two leukocyte telomere-length datasets [35] [36] [37]. Inflammatory sources comprised primary and alternative CRP, circulating IL-6 and GlycA [38] [39] [40] [41]. Source GWAS scales were retained. SGS and UK Biobank contributed to multiple sources; exact overlap covariances were unavailable. Accessions, population definitions and unavailable models are listed in Supplementary Table S1 and Methods S9.

Instruments met P < 5 × 10⁻⁸ and were clumped at r² < 0.001 within 10 Mb using a European reference. Associations were aligned on GRCh37 without LD proxies. Palindromic variants were excluded in Ruth-connected analyses; other pairs used TwoSampleMR action-2 harmonization. Primary MR used multiplicative random-effects IVW, with variance floored at one [42]. MR-Egger, weighted median and weighted mode estimates provided sensitivity analyses where at least three variants were available [43] [44] [45]. Computation used frozen TwoSampleMR 0.6.22 functions [46]. Diagnostics included heterogeneity, the Egger intercept, leave-one-out analysis and instrument strength. These analyses assume relevant instruments without confounding or outcome pathways that bypass the instrumented exposure.

### Blood-cell and inflammatory-pathway analyses

Measured neutrophil percentage, neutrophil count and lymphocyte percentage came from sex-specific and pooled Neale Lab UK Biobank GWAS. Male comparisons used female SNP–ANM coefficients as the exposure scale, following a menopause genetic negative-control design [16]. Female–male contrasts assumed independent outcome errors conditional on shared exposure coefficients, without overlap or exposure-estimation covariance correction, and retained separately rank-normalized outcome scales. These were blood-cell association analyses, separate from mediation.

Multivariable IVW jointly modelled ANM and each inflammatory biomarker using the reclumped union of aligned instruments. Conditional F < 10 flagged weak instruments, assuming zero cross-exposure sampling covariance [47] [24]. Indirect effects were a × b: the primary IVW ANM-to-biomarker coefficient multiplied by the biomarker coefficient conditional on ANM. Both paths used the same biomarker source; first-order delta variance assumed zero path covariance [48]. Interpretation requires valid conditional instruments, compatible scales and causal assumptions; exposure–outcome sex differences and shared samples remain relevant. Formulas, component estimates and full assumptions are in Supplementary Methods S9–S10.

### Multiple testing and reproducibility

The main observational correction family comprised all thirteen markers across both cohorts (26 tests), including the supporting PC analyses. All thirteen markers were included in the joint imputation. Genetic correction retained the original planned families separately by direction, estimator and coefficient role; multivariable ANM, biomarker and indirect coefficients each used a separate 50-test family, and sex contrasts formed a three-test family. Unavailable tests contributed P = 1 for adjustment, without assigned estimates. Complete correction inventories are in Supplementary Methods S11. Effect estimates, uncertainty, P values and q values are provided in the source data. Figures were produced in R.

## Data availability

Generation Scotland participant-level records are controlled-access data available through the cohort’s governance procedures [27]. Supplementary Data 1 contains the aggregate cohort results and diagnostics; Supplementary Data 2 contains genetic estimates and diagnostics; Supplementary Data 3 contains available instrument-level and sensitivity evidence. The Source Data workbook and figure-specific CSVs provide the displayed values. Provider-approved access wording and permanent deposit details remain for author completion.

All data produced in the present study are available upon reasonable request to the authors

## Code availability

The code used in this study is available upon request.

## Acknowledgments

Cohort acknowledgments, funding and grant details are to be completed by the authors.

